# Higher FSH Level Is Associated With Increased Risk Of Incident Hip Fracture In Older Adults, Independent Of Sex Hormone Levels

**DOI:** 10.1101/2024.02.19.24303043

**Authors:** Eileen H. Koh, Susan K. Ewing, Sigurdur Sigurdsson, Vilmundur Gudnason, Trisha F. Hue, Eric Vittinghoff, Claes Ohlsson, Åsa Tivesten, Louise Grahnemo, Tony Yuen, Mone Zaidi, Clifford J. Rosen, Ann V. Schwartz, Anne L. Schafer

## Abstract

**Context:** Higher levels of follicle stimulating hormone (FSH) are associated with bone loss among women during the perimenopausal transition and among older men, independent of estradiol and testosterone levels, but it is unknown whether higher FSH is an independent risk factor for fracture.

**Objective:** Determine whether baseline FSH level predicts subsequent hip fracture in older adults.

**Setting, Design, Participants:** Using a case-cohort design, we randomly sampled 295 participants stratified by sex from the Age, Gene/Environment Susceptibility (AGES)-Reykjavik cohort, including 25 participants with incident hip fracture within 10 years after baseline. We sampled an additional 230 sex-stratified participants with incident hip fracture. Serum FSH and sex hormone levels were measured at baseline. Robust weighted Cox proportional hazards models were used to determine the relationship between FSH and hip fracture risk.

**Main Outcome:** Incident hip fracture

**Results:** As no interaction was identified between FSH and sex for the relationship with fracture, men and women were pooled for analysis. Higher levels of FSH were associated with a significantly increased risk of incident hip fracture in models adjusted for age and sex [hazard ratio (HR) 1.24 (95% CI 1.04-1.48, p=0.02)] and after further adjustment for estradiol, testosterone, and sex hormone binding globulin levels [HR 1.20 (95% CI 1.01-1.44, p=0.04) per sex-specific SD increase in FSH level].

**Conclusions:** Higher FSH is associated with increased risk of subsequent hip fracture. Our findings support a growing body of evidence for direct pleiotropic effects of FSH on bone, and for a role for FSH in aging and disability independent of sex hormone levels.

## INTRODUCTION

The relationship between follicle stimulating hormone (FSH) and bone was traditionally attributed to FSH’s regulation of sex hormones. However, emerging evidence suggests that FSH directly impacts the skeleton, independently of sex hormone levels. In the Study of Women’s Health Across the Nation (SWAN), higher FSH levels were associated with higher concentrations of bone turnover markers and lower bone mineral density (BMD) in premenopausal and early perimenopausal women, whereas estradiol, testosterone, and sex hormone binding globulin (SHBG) were not (1,2). Furthermore, in this cohort, during the menopausal transition when FSH levels rise but estradiol levels remain stable, higher baseline and subsequent follow-up FSH levels—but not estradiol, testosterone, or SHBG levels—were predictive of greater reductions in BMD at spine and hip (3). Among men, in a cross-sectional case-control study of middle-aged men with osteoporosis and healthy controls, FSH was negatively associated with BMD even after adjustment for testosterone and SHGB levels (4).

The relationship between FSH and bone has also been studied in older adults. Cross-sectionally, among postmenopausal women (mean age 81 years) in the Age Gene/Environment Susceptibility (AGES)-Reykjavik cohort study, we found that higher FSH levels were associated with lower bone density and decreased bone strength independent of estradiol and testosterone levels (5). This relationship was not seen among men. In contrast, in the Concord Health and Aging in Men Project (CHAMP) prospective study of aging men >70 years of age, FSH levels predicted hip BMD loss while testosterone levels did not. Each 1 standard deviation (SD) decrease in FSH was associated with a 21% reduction in incident hip fracture risk, though this finding was no longer significant after adjusting for age, BMI and other confounders (6).

Direct effects of FSH on the skeleton were first documented through genetic and pharmacological studies in mice. Mice haploinsufficient in *Fshr,* the gene encoding the FSH receptor, were found to have high bone mass with conserved ovarian function, in essence separating the effects of FSH and estrogen on bone (7). FSH was further found to increase bone resorption and reduce bone formation, and as a result to cause net bone loss in a number of animal models (8,9). We and others found that blocking FSH action using antibodies designed to prevent the interaction of FSHβ, or an FSHβ fusion antigen given as a vaccine, prevented ovariectomy-induced bone loss by inhibiting bone resorption and stimulating bone formation (10–16).

Taken together, the preclinical models and human observational studies suggest that FSH regulates bone mass independent of sex hormone levels. However, it remains unclear whether the negative effect on bone mass translates to increased fractures, in particular among older adults who are at greatest risk for fracture. Using the Age Gene/Environment Susceptibility (AGES)-Reykjavik cohort, we hypothesized that higher levels of FSH would predict increased risk of incident hip fracture among older adults, independent of sex hormone levels.

## MATERIALS AND METHODS

### Participants and Study Design

The Age Gene/Environment Susceptibility (AGES)-Reykjavik Study is a longitudinal, observational, population-based study of older adults living in Iceland, designed to assess gene/environment interactions and their relationships to diseases and disability in old age (17). Participants were randomly selected from surviving members of the Reykjavik study, a cohort established in 1967 to study cardiovascular risk factors (18,19). A total of 5,764 older adults aged 66 to 96 participated in the baseline AGES-Reykjavik visit between 2002 and 2006. The AGES-Reykjavik Study has been approved by the Icelandic National Bioethics Committee and by the Institutional Review Board for the Intramural Research Program of the National Institute on Aging, National Institutes of Health. Written informed consent was obtained from all participants.

For this analysis of the relationship between serum FSH level and hip fracture risk, 58 participants were excluded for not granting access to hospital records. Participants were also excluded if they had a history of hip fracture prior to AGES baseline (N=16 women, N=12 men); if quantitative computed tomography (QCT) scans for spine and hip were unavailable (N=622 women, N=321 men); or if they reported use of certain medications known to affect FSH, sex hormone levels or bone mineral density (N=634 women, N=124 men). Exclusionary medications were sex hormone replacement therapy including SERMs (N=468 women, N=8 men), anti-androgens (N=44 men), anti-estrogens (N=45 women), aromatase inhibitors (N=7 women), GnRH agonists (N=3 men), and systemic glucocorticoids (N=140 women, N=72 men). After applying exclusions, 4,144 participants (72% of participants at AGES baseline, 2,152 women and 1,992 men) were eligible for the analysis (Figure). Among those eligible, 380 participants (255 women and 125 men) sustained an incident hip fracture within 10 years of the baseline visit.

**Figure.**
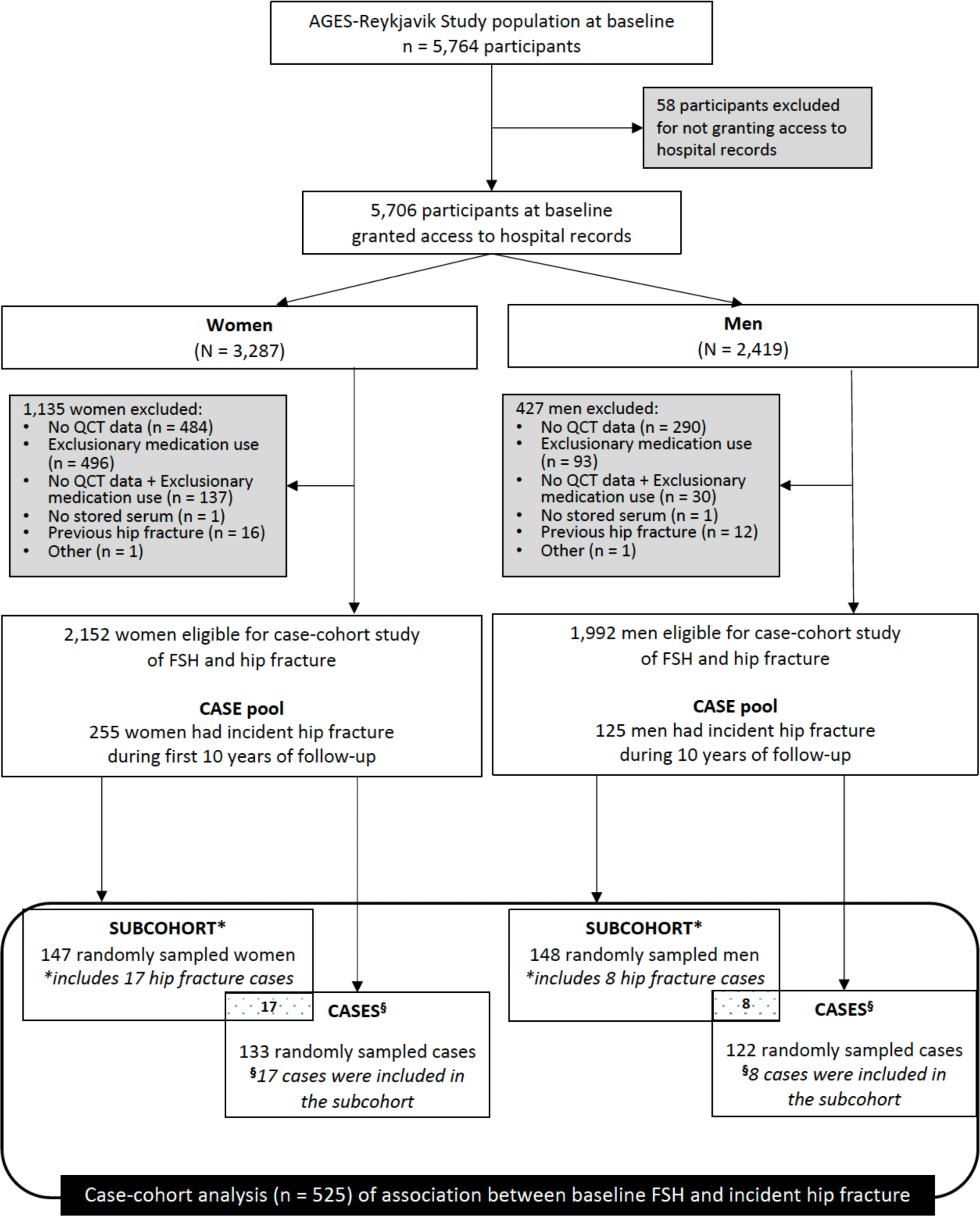
Legend to Figure. Approach to sampling for the case-cohort study. QCT, quantitative computed tomography. FSH, follicle stimulating hormone.

A case-cohort design was used to examine the relationship between serum FSH and incident hip fracture risk. Our subcohort of 295 participants (147 women and 148 men) was randomly sampled, stratified by sex, from the pool of 4,144 eligible participants. The random subcohort included 25 incident hip fracture cases. An additional 230 hip fracture cases were randomly sampled, stratified by sex, from the remaining case pool of 355 eligible participants. Hip fracture cases thus comprised 255 participants (133 women and 122 men) (Figure). Power calculations used in the design of the study indicated that for a random subcohort of 300 and 100 hip fracture cases for each sex, we would have 80% power to detect a HR=1.30 per SD change in FSH for the pooled analysis and HR=1.45 for each sex. In implementing the study, we added more hip fracture cases and thus improved statistical power, lowering the detectable HR.

### Sex Hormone Measurements

At the AGES baseline visit, venous blood samples were collected between 8:00 and 11:30 AM after an overnight fast and stored at −80 °C. For this case-cohort study, archived serum specimens from sampled participants were batch analyzed for FSH, sex hormone levels, and SHBG.

Serum FSH level was measured at MaineHealth Institute for Research (Scarborough, ME) as a single batch using ELISA (ALPCO, Salem, NH, USA). The assay sensitivity was 1 IU/L and there was no cross-reactivity with high levels of hCG (1000-50,000IU/L), LH (5-250 IU/L) or TSH (5-250 mIU/L). Intra-assay coefficient of variation (CV) was 4.2%, and inter-assay CV was 5.2%.

The sex steroids total estradiol and total testosterone (to be referred to as “estradiol” and “testosterone”) were measured at Sahlgrenska Academy (Gothenburg, Sweden). Sera from both men and women were batch analyzed by liquid chromatography-tandem mass spectrometry (LC-MS/MS). The lower limit of quantification (LLOQ) for estradiol is 0.5 pg/mL (0.05 ng/dL) and testosterone 5 pg/mL (0.5 ng/dL). For estradiol, the intra-assay CV was 1.1-2.3% and inter-assay CV 1.1-1.9%. For testosterone, the intra-assay CV was 1.1-1.9% and inter-assay CV 2.2-2.4% (20).

Serum SHBG in women was batch analyzed at University of Turku by a chemiluminescent-based SHBG assay (LIAISON SHBG, cat. code 319020, DiaSorin; RRID:AB_2895155). The LLOQ of the assay is 0.80 nmol/L. Intra-assay CV is typically <3% and inter-assay CV <6%.

Serum SHBG in men was batch analyzed at MaineHealth Institute for Research using ELISA (R&D Systems). Lower limit of detection is 0.005 nmol/L (sensitivity 0.012 nmol/L) with intra-assay CV 4.1% and inter-assay CV 6.1%.

### Measures of Bone Mineral Density

Quantitative computed tomography (QCT) for bone mineral density was performed in the lumbar spine (L1 and L2 vertebrae) and hip using a 4-detector CT system (Sensation, Siemens Medical Systems, Erlangen, Germany). To calibrate CT Hounsfield units to equivalent concentration of hydroxyapatite, a bone mineral reference standard (3-sample calibration phantom, Image Analysis, Columbia, KY) was placed under the participant’s spine and hips and scanned simultaneously. The lumbar spine scanning included a helical study of the L1 and L2 vertebrae (120 kVp, 150 mAs 1-mm slice thickness, pitch = 1). A helical study of the hip (120 kVp, 140 mAs, 1-mm slice thickness, pitch = 1) included the proximal femur from a point 1 cm superior to the acetabulum to a point 3–5 mm inferior to the lesser trochanter. (21). QCT images were processed to extract measures of volumetric BMD (vBMD) in g/cm^3^ using analytical algorithms (22). Scanner stability was monitored using reproducible daily quality assurance tests based on a phantom test, including measurements of slice geometry, spatial uniformity, density linearity, spatial resolution, and noise. The imaging center also performed weekly measurements to monitor density linearity of the calibration phantom described above and calibrated the scanner monthly against water.

### Incident Fracture Ascertainment

Fracture data were recorded, verified, and confirmed using medical and radiologic records within the AGES-Reykjavik fracture registry, which has previously been described (23,24). Hip fractures were defined by ICD-10 diagnostic codes (S72.0, S72.1, S72.2). The fracture registry has been shown to have a hip fracture capture rate of 97%. Fractures with avulsion detachments <5×6 mm^2^, pathological fractures due to malignancy, and stress fractures were excluded. Furthermore, any fracture occurring between the date of entry into the Reykjavik Study in 1967 and the date of entry into the AGES-Reykjavik Study 2002-2006 was also recorded, from which we were able to identify participants with a history of hip fracture prior to AGES baseline.

### Other Measures

The AGES baseline visit included blood samples, anthropometry (height and weight), and standardized questionnaires addressing topics including demographics, health history, and medication use (17). Diabetes was defined by self-report, diabetes medication use, fasting plasma glucose ≥7 mmol/L, or hemoglobin A1c (HbA1c) ≥6.5%. Body mass index (kg/m^2^) was calculated based on height and weight. Estimated glomerular filtration rate (eGFR) was calculated using the Chronic Kidney Disease Epidemiology Collaboration equation.

### Statistical Analysis

Baseline characteristics of the random subcohort and hip fracture cases were summarized using means and SD for continuous variables, and counts and percentages for categorical variables. Mean baseline FSH and sex hormone levels of the random subcohort and hip fracture cases were reported separately for women and men. High values of estradiol, testosterone, and SHBG were set to the highest observed value below the sex-specific mean + 2 SD; the random subcohort was used to determine the cutoffs.

To assess the independent association of baseline FSH level with incident hip fracture during the first 10 years of follow-up, we used Cox proportional hazards models with the Barlow weighting method and robust variance estimation to accommodate the sex-stratified sampling and case-cohort design. According to Barlow’s method for weighting participants in the pseudolikelihood, fracture cases were weighted by 1 and subcohort controls were weighted by the inverse of the sampling fraction (alpha = n/N where n is the number of women or men in the random subcohort, and N is the total number of women or men at AGES baseline, after applying exclusions): alpha = 147/2152 = 6.8% for women and alpha = 148/1992 = 7.4% for men. Time to event in the models was set to time to first hip fracture for cases, time to death for subcohort controls who died within the first 10 years of follow-up, and 10 years for those in the subcohort who did not die within the follow-up period. We expressed FSH as a continuous variable in our analysis, as tests for non-linearity using quartiles of FSH were negative. The risk of incident hip fracture per sex-specific SD increase in FSH was estimated for each sex and overall, with results presented as hazard ratios (HR) and 95% confidence intervals (CIs). Unadjusted and age-adjusted Cox regression was performed, as well as models adjusted for age, sex (pooled model only), estradiol, testosterone, and SHBG. We determined whether FSH was associated with hip fracture risk independently of vBMD. The interaction between baseline FSH and sex was tested to determine if associations between FSH and incident hip fracture differed for women and men. Two sensitivity analyses were performed: (1) with baseline bisphosphonate users excluded from the models and (2) with further adjustment for BMI, diabetes or eGFR in the models. All analyses were performed with SAS software (version 9.4, SAS Institute Inc., Cary, NC, USA).

## RESULTS

### Baseline Characteristics and Follow-up Time

The characteristics of the random subcohort (N=295) and hip fracture cases (N=255) are provided in Table 1. The mean age of the random subcohort was 75.9 ± 5.1 years, and their mean BMI was 27.1 ± 4.2 kg/m^2^. Among the random subcohort, mean follow-up time was 8.7 ± 2.4 years, and 34% died during the 10-year follow-up. Among cases, mean time to incident hip fracture was 5.7 ± 2.5 years.

**Table 1:** Baseline characteristics of random subcohort and hip fracture cases.

|  |  |  |
| --- | --- | --- |
|  | Subcohort*<br>(n =295) | Hip Fracture Cases (n = 255) |
| Age (years) | 75.9 ± 5.1 | 79.2 ± 5.3 |
| Women | 147 (49.8) | 133 (52.2) |
| Age of menopause (years, women only) | 47.6 ± 4.9 | 47.9 ± 4.8 |
| BMI (kg/m<sup>2</sup>) | 27.1 ± 4.2 | 26.1 ± 4.4 |
| Diabetes | 41 (13.9) | 34 (13.3) |
| Congestive Heart Failure | 15 (5.1) | 16 (6.3) |
| Stroke | 19 (6.5) | 22 (8.7) |
| Cancer | 40 (13.6) | 38 (15.0) |
| Chronic Obstructive Pulmonary Disease | 24 (8.3) | 26 (10.3) |
| Active smoker | 40 (13.6) | 29 (11.4) |
| Former smoker | 146 (49.5) | 114 (44.7) |
| History of prior fracture | 155 (52.5) | 150 (58.8) |
| History of osteoporosis | 33 (11.4) | 31 (12.8) |
| History of fall in prior 12 months | 47 (16.0) | 57 (22.4) |
| Use of osteoporosis medications** | 11 (3.7) | 7 (2.8) |
| eGFR (mL/min/1.73 m<sup>2</sup>) | 64.3 ± 15.7 | 61.7 ± 16.1 |
| **Bone Mineral Density** |  |  |
| Total Hip integral vBMD (g/cm<sup>3</sup>) | 0.237 ± 0.046 | 0.206 ± 0.040 |
| Femoral Neck integral vBMD (g/cm<sup>3</sup>) | 0.246 ± 0.049 | 0.214 ± 0.041 |
| Spine integral vBMD (g/cm<sup>3</sup>) | 0.113 ± 0.035 | 0.098 ± 0.034 |
| Data in table are presented as mean ± SD or n (%). |  |  |
| eGFR was calculated with the CKD-EPI formula. |  |  |
| * 25 participants in the incident hip fracture cases were also randomly selected as part of the subcohort |  |  |
| ** Medications included bisphosphonates and calcitonin |  |  |

### Baseline Serum FSH and Sex Hormone Levels

Among women in the random subcohort, mean FSH was 64.3 ± 25.2 IU/L with estradiol 5.7 ± 4.2 pg/mL, testosterone 22.5 ± 10.4 ng/dL, and SHBG 74.0 ± 33.8 nmol/L (Table 2). Among women hip fracture cases, mean FSH was higher at 69.9 ± 27.4 IU/L, estradiol was slightly lower, while testosterone and SHBG were higher. The mean age of menopause was similar between groups (Table 1). Two women in the subcohort had FSH values <18 IU/L. Because they had expected postmenopausal range estradiol levels, they were included in our analysis for generalizability. Among men in the subcohort, mean FSH was 12.2 ± 11.9 IU/L with estradiol 21.3 ± 6.9 pg/mL, testosterone 457.9 ± 171.2 ng/dL, and SHBG 52.8 ± 17.5 nmol/L. Among male hip fracture cases, baseline FSH was higher, mean estradiol and testosterone were slightly lower, and SHBG was higher.

**Table 2:**
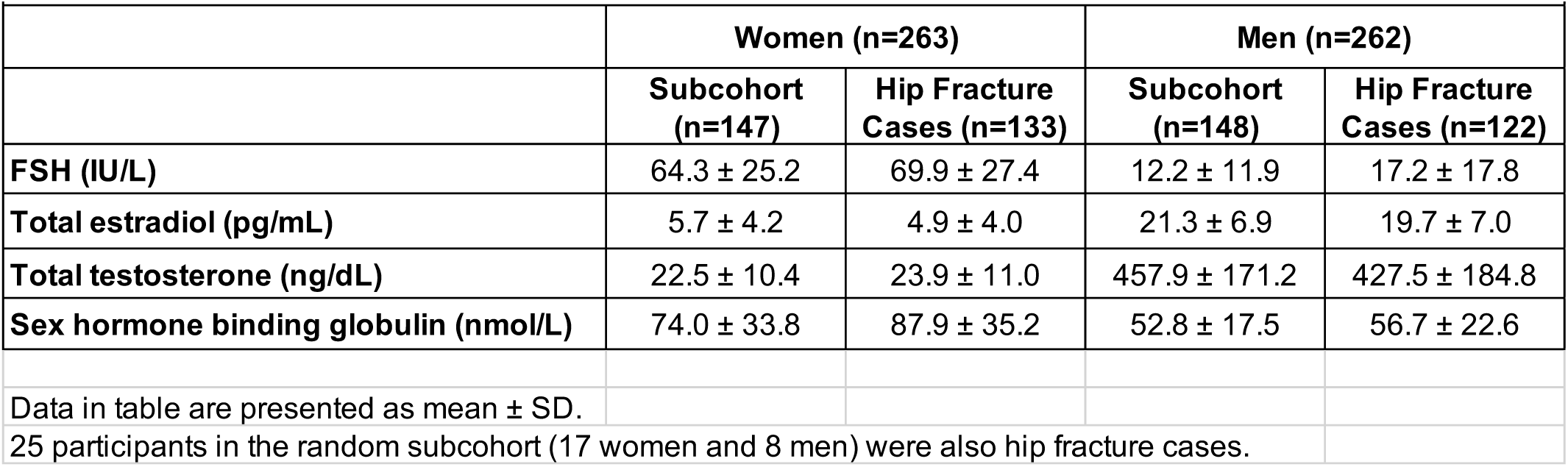
FSH and sex hormone levels of random subcohort and hip fracture cases stratified by sex.

### Association Between Baseline FSH and Incident Hip Fracture

Among all participants, the risk of incident hip fracture increased 24% per sex-specific SD increase in FSH level in models adjusted for age and sex [hazard ratio (HR) 1.24 (95% CI 1.04-1.48), p=0.02)] (Table 3). Further adjustment for estradiol, testosterone, and SHBG levels attenuated this risk slightly to 20%, but the risk remained statistically significant [HR 1.20 (95% CI 1.01-1.44), p=0.04]. In models adjusted for age, estradiol, testosterone, and SHBG, each SD increase in FSH level was associated with a 22% increased risk of hip fracture among women [HR 1.22 (95% CI 0.92-1.61), p=0.16] and 19% increased risk among men [HR 1.19 (95% 0.93-1.52), p=0.16]. There was no interaction between FSH and sex in the models (all interaction p-values ≥0.83), thus women and men were pooled for primary analyses.

**Table 3:** Risk of incident hip fracture per sex-specific SD increase in FSH, stratified by sex and pooled.

|  |  |  |  |  |
| --- | --- | --- | --- | --- |
|  | **All participants\*** | **Women** | **Men** | **FSHxSex**<br>**interaction** |
|  | **HR (95% CI)** | **HR (95% CI)** | **HR (95% CI)** | **p-value** |
| Model 1 (unadjusted) | 1.28 (1.08, 1.52)** | 1.27 (0.99, 1.62) | 1.29 (1.03, 1.61)** | 0.93 |
| Model 2 (adjusted for age) | 1.24 (1.04, 1.48)** | 1.25 (0.97, 1.62) | 1.23 (0.96, 1.56) | 0.90 |
| Model 3 (adjusted for age, estradiol, testosterone, and SHBG) | 1.20 (1.01, 1.44)** | 1.22 (0.92, 1.61) | 1.19 (0.93, 1.52) | 0.83 |
| Sex-specific SD for FSH determined from random subcohort (SD=25.2 IU/L for women, SD=11.9 IU/L for men) |  |  |  |  |
| * Models for all participants were also adjusted for sex |  |  |  |  |
| ** denotes p-value < 0.05 |  |  |  |  |

We did not find evidence of a cross-sectional association between FSH and vBMD. For each SD increase in FSH, mean difference in total hip integral vBMD was -0.0006 g/cm^3^ (p=0.71), mean difference in femoral neck vBMD was -0.0011 g/cm^3^ (p=0.52), and mean difference in spine integral vBMD was -0.0012 g/cm^3^ (p=0.35). Adding integral vBMD did not substantially change the relationship between FSH and hip fracture risk: Adjusted HR was 1.20 (p=0.04) before addition of vBMD, then HR was 1.21 (p=0.052) with addition of total hip vBMD; HR was 1.23 (p=0.04) with femoral neck vBMD; and HR was 1.19 (p=0.06) with spine vBMD. These findings indicate that baseline integral vBMD is not an important mediator of the relationship between FSH and hip fracture risk.

There was no statistically significant association between FSH and BMI (beta=-0.20 per SD increase in FSH, p=0.16), nor between BMI and hip fracture risk (HR 1.05 per 1 kg/m^2^ decrease in BMI, p=0.12). Adding BMI, diabetes status, and eGFR separately to the models adjusted for age, sex, estradiol, testosterone, and SHBG had little or no effect on the association between FSH and hip fracture risk (HR=1.20). With the addition of BMI, diabetes status, or eGFR, the respective associations between FSH and incident fracture were HR 1.18 (95% CI 0.99-1.42), HR 1.20 (95% CI 1.01-1.44), and HR 1.18 (95% CI 0.98-1.41).

In a sensitivity analysis excluding 18 individuals (17 women and 1 man) who reported use of osteoporosis medications at baseline, the estimated hazard ratio for the association between FSH and hip fracture risk was not altered, although it was no longer statistically significant [HR 1.20 (95% CI 1.00-1.44), p=0.055 per SD increase in FSH, after adjustment for age, sex, estradiol, testosterone, and SHBG].

## DISCUSSION

This is the first study to investigate the relationship between serum FSH level and risk of hip fracture in older men and women, independent of estradiol and testosterone levels. We report that higher serum FSH level was associated with increased risk of incident hip fracture, with a risk increase of 20% per sex-specific SD increase in FSH level after adjusting for age, sex, sex hormone levels, and SHBG.

Although a body of literature now links FSH and bone, few have investigated whether FSH level predicts subsequent fracture, and ours is the first study to our knowledge to have demonstrated a relationship between FSH level and incident fracture risk independent of sex hormone levels. This could suggest several potential clinical applications to reduce overall fracture morbidity and mortality. Measuring serum FSH level may assist in identifying, among older adults, those who are at highest risk of hip fracture. Furthermore, there may be a role for therapeutic FSH blockade to prevent fractures in humans, such as with a humanized antibody (11,12), as FSH blockade in rodent models has been shown to have osteoprotective effects (13,15,25). We plan to move to clinical trials a first-in-class humanized FSH-blocking antibody (11,12), not only for the outcome of osteoporosis but also for obesity and cognitive decline (14,26), other conditions that track with bone loss across the menopausal transition (27–29).

Prior studies have not identified an association between FSH and incident fracture. Although in SWAN higher baseline FSH and subsequent FSH level was predictive of 4-year areal BMD loss at the lumbar spine in women undergoing menopausal transition (3), in a subsequent SWAN study FSH level did not correlate with self-reported incident fracture (relative risk = 1.06; 95% CI 0.95-1.17) over 8.8 years of follow-up with cumulative incident fracture rate of 17% (30). We reason that fracture risk may not have been high enough in the early menopausal women to have detected a relationship between FSH and fracture, or that elevated FSH levels in perimenopausal women may not have had time to impart substantive changes to bone to increase fracture risk. In comparison, the women in our study were on average 28 years postmenopausal at time of baseline visit. Our study also benefits from a particularly robust ascertainment of fracture, as all incident fractures were adjudicated.

In the CHAMP study of older men, higher baseline FSH level was associated with greater bone loss at the hip (as were higher LH and SHBG and lower estrone), while no statistically significant association was found between testosterone or estradiol and change in hip BMD. While in their unadjusted models, one SD decrease in FSH level was associated with a 13% reduction in any fracture and a 21% reduction in risk of hip fracture, this association was no longer significant after adjusting for age and other factors (HR for any fracture 0.93 (95% CI 0.82–1.07) and HR for hip fracture 0.93 (95% CI 0.73–1.18) (6). The negative CHAMP study findings with respect to the association of FSH and hip fracture risk might be explained by a 45% rate of loss to follow-up at 5-years (predominantly due to death), and an overall low rate of incident hip fractures (3%).

In our analysis, the cross-sectional association between FSH level and hip vBMD in older women and men was in the negative direction, but was not statistically significant. Previous studies have reported a statistically significant negative association for women, including our own cross-sectional study in a subset of AGES-Reykjavik participants at a different (later) time point (5). As described above, in SWAN, among a multiethnic group of 2335 pre- and perimenopausal women (age 42-52 years), FSH was strongly correlated with lower areal BMD at lumbar spine and hip, while estradiol and testosterone were not (1). In a retrospective study of women taking menopausal hormone therapy, change in FSH correlated with change in BMD after 12 and 24 months of therapy (31). Other studies have reported a lack of association between FSH and BMD in men, including our own cross-sectional AGES-Reykjavik study at the later time point (5). One study of middle-aged men with infertility reported that 15 years after infertility diagnosis, BMD measurements were no different when compared to age-matched healthy men despite higher median FSH levels (9.8 v 3.7 IU/L) (32). In a similar cohort involving 307 men with idiopathic infertility (mean FSH 5.3 IU/L) and 28 men with Klinefelter syndrome (mean FSH 35.7 IU/L), serum FSH levels did not significantly correlate with BMD (33).

Our findings indicate that vBMD is unlikely an important mediator of the relationship between FSH and hip fracture. This suggests that FSH might have detrimental effects on bone strength beyond its effects on bone mass as measured by vBMD, such as effects on bone quality. Furthermore, FSH may also increase fracture risk via its effects on non-bone targets, such as skeletal muscle, visceral adiposity, or cognition that may increase fragility and risk of falls (5,34–36). Preclinical models have shown that FSH worsens cognitive decline in mice prone to Alzheimer’s disease, and these effects were prevented by FSH blockade (26).

One might expect a participant’s sex to modulate the relationship of FSH and hip fracture, given the studies described above that have demonstrated an association between FSH and BMD in older women but not men. However, we did not find an interaction between sex and FSH for the risk of hip fracture, suggesting that high levels of circulating FSH confer similarly elevated relative risk for hip fracture in older men and women. It also impresses the importance of including older men in future studies examining the relationship between FSH and osteoporosis.

A major strength of our study was use of a well-characterized longitudinal cohort, which enabled us to examine the temporal relationship between FSH and incident hip fracture risk. Our hip fracture outcomes were identified from an adjudicated fracture registry with a hip fracture capture rate of 97%. Another strength was the use of mass spectrometry for sex hormone measurements. Mass spectrometry allows for accurate quantification of lower levels of circulating sex steroids, which is particularly relevant in men (37) and for our older adult cohort.

Limitations of our study include lack of baseline bone turnover marker levels and DXA assessment of bone or body composition, and lack of follow-up measurement of FSH and sex hormone measurements. Although we controlled for age, sex, and sex hormone levels, and we carefully examined other candidate covariates for potential inclusion in our models, there may be other, unmeasured confounders which might account for the observed association. That our study was conducted in a predominantly white population from a single country limits generalizability of our findings.

In conclusion, our case-cohort study of older healthy adults from Iceland demonstrates that higher serum FSH is associated with increased risk of hip fracture, independent of sex hormone levels. Further studies are needed to better understand the biological mechanisms behind our findings.

## Data Availability

All data produced in the present study are available through collaboration under a data usage agreement with the Icelandic Heart Association. (Based on the informed consent provided by AGES-Reykjavik participants, analyses using the data can only be carried out after approval from the Icelandic data protection authority.)

## Funding

This study was funded by the National Institute on Aging (NIA, U19AG060917), with additional support from the National Institute of Arthritis and Musculoskeletal and Skin Diseases (NIAMS, R01AR057819, R01AR065645, P30AR075055). The AGES-Reykjavik Study is supported by funding from the NIA (N01AG12100), the NIA Intramural Research program, Hjartavernd (the Icelandic Heart Association), and the Althingi (Icelandic Parliament). EHK has received support from the National Institute of Diabetes and Digestive and Kidney Diseases (NIDDK, T32DK007418).

## Competing interests

All authors have completed the ICMJE uniform disclosure form at www.icmje.org/coi_disclosure.pdf and declare: EHK, SKE, SS, VG, TFH, EV, TY, MZ, CJR, AVS, and ALS had support from the National Institutes of Health for the submitted work. SS and VG also had support from Hjartavernd (Icelandic Heart Association) and the Althingi (Icelandic Parliament) for the submitted work. CO has received research support from the Swedish Research Council and grants from the Swedish state under the agreement between the Swedish government and the county councils, the ALF agreement, from the Novo Nordisk Foundation, and from the Knut and Alice Wallenberg Foundation; he also has two patents/ patent applications in the field of probiotics and bone health. MZ has patents addressing FSH, FSH formulation, bone, body fat, and neurodegeneration, and MZ and TY have patents addressing luteinizing hormone (LH) and body composition. ALS has received investigator-initiated research grant support from Amgen addressing bone health after bariatric surgery, and grant support from Bone Health Technologies. There are no other relationships or activities that could appear to have influenced the submitted work.

## Data availability and sharing

Data from the AGES-Reykjavik Study are available through collaboration under a data usage agreement with the Icelandic Heart Association.

## Notes

### Author Declarations

The Icelandic National Bioethics Committee and the Institutional Review Board of the Intramural Research Program of the National Institute on Aging, National Institutes of Health gave ethical approval for this work.

## REFERENCES

1. Sowers MR, Finkelstein JS, Ettinger B, et al. The association of endogenous hormone concentrations and bone mineral density measures in pre- and perimenopausal women of four ethnic groups: SWAN. Osteoporos Int. 2003; 14:44–52.

2. Sowers MR, Greendale GA, Bondarenko I, et al. Endogenous hormones and bone turnover markers in pre- and perimenopausal women: SWAN. Osteoporos Int. 2003; 14:191–197.

3. Sowers MR, Jannausch M, McConnell D, et al. Hormone predictors of bone mineral density changes during the menopausal transition. J Clin Endocrinol Metab. 2006; 91:1261–1267.

4. Karim N, MacDonald D, Dolan AL, Fogelman I, Wierzbicki AS, Hampson G. The relationship between gonadotrophins, gonadal hormones and bone mass in men. Clin Endocrinol (Oxf). 2008; 68:94–101.

5. Veldhuis-Vlug AG, Woods GN, Sigurdsson S, et al. Serum FSH is associated with BMD, bone marrow adiposity, and body composition in the AGES-Reykjavik study of older adults. J Clin Endocrinol Metab. 2021; 106:e1156–e1169.

6. Hsu B, Cumming RG, Seibel MJ, et al. Reproductive hormones and longitudinal change in bone mineral density and incident fracture risk in older men: The Concord Health and Aging in Men Project. J Bone Miner Res. 2015; 30:1701–1708.

7. Sun L, Peng Y, Sharrow AC, et al. FSH directly regulates bone mass. Cell. 2006; 125:247–260.

8. Zaidi M, Yuen T, Kim SM. Pituitary crosstalk with bone, adipose tissue and brain. Nat Rev Endocrinol. 2023;

9. Liu S, Cheng Y, Fan M, Chen D, Bian Z. FSH aggravates periodontitis-related bone loss in ovariectomized rats. J Dent Res. 2010; 89:366–371.

10. Geng W, Yan X, Du H, Cui J, Li L, Chen F. Immunization with FSHbeta fusion protein antigen prevents bone loss in a rat ovariectomy-induced osteoporosis model. Biochem Biophys Res Commun. 2013; 434:280–286.

11. Gera S, Kuo TC, Gumerova AA, et al. FSH-blocking therapeutic for osteoporosis. Elife. 2022; 11:

12. Gera S, Sant D, Haider S, et al. First-in-class humanized FSH blocking antibody targets bone and fat. Proc Natl Acad Sci U S A. 2020; 117:28971–28979.

13. Ji Y, Liu P, Yuen T, et al. Epitope-specific monoclonal antibodies to FSHbeta increase bone mass. Proc Natl Acad Sci U S A. 2018; 115:2192–2197.

14. Liu P, Ji Y, Yuen T, et al. Blocking FSH induces thermogenic adipose tissue and reduces body fat. Nature. 2017; 546:107–112.

15. Zhu LL, Blair H, Cao J, et al. Blocking antibody to the beta-subunit of FSH prevents bone loss by inhibiting bone resorption and stimulating bone synthesis. Proc Natl Acad Sci U S A. 2012; 109:14574–14579.

16. Zhu LL, Tourkova I, Yuen T, et al. Blocking FSH action attenuates osteoclastogenesis. Biochem Biophys Res Commun. 2012; 422:54–58.

17. Harris TB, Launer LJ, Eiriksdottir G, et al. Age, Gene/Environment Susceptibility-Reykjavik Study: Multidisciplinary applied phenomics. Am J Epidemiol. 2007; 165:1076–1087.

18. Bjornsson G BO, Davidsson D 1982 Report abc XXIV Health survey in the Reykjavik area - women stages i-iii 1968-1969, 1971-1972, and 1976-1978. Participants invitation response etc. Reykjavik: Icelandic Heart Association.

19. Bjornsson O DD, Olafsson H 1979 Report abc XVIII Health survey in the Reykjavik area - men stages i-iii 1967 - 1969, 1970-1971 and 1974-1976. Participants invitation response etc. Reykjavik: Icelandic Heart Association.

20. Ohlsson C, Langenskiold M, Smidfelt K, et al. Low progesterone and low estradiol levels associate with abdominal aortic aneurysms in men. J Clin Endocrinol Metab. 2022; 107:e1413–e1425.

21. Sigurdsson G, Aspelund T, Chang M, et al. Increasing sex difference in bone strength in old age: The Age, Gene/Environment Susceptibility-Reykjavik study (AGES-Reykjavik). Bone. 2006; 39:644–651.

22. Lang T, LeBlanc A, Evans H, Lu Y, Genant H, Yu A. Cortical and trabecular bone mineral loss from the spine and hip in long-duration spaceflight. J Bone Miner Res. 2004; 19:1006–1012.

23. Siggeirsdottir K, Aspelund T, Sigurdsson G, et al. Inaccuracy in self-report of fractures may underestimate association with health outcomes when compared with medical record based fracture registry. Eur J Epidemiol. 2007; 22:631–639.

24. Marques EA, Gudnason V, Lang T, et al. Association of bone turnover markers with volumetric bone loss, periosteal apposition, and fracture risk in older men and women: The AGES-Reykjavik longitudinal study. Osteoporos Int. 2016; 27:3485–3494.

25. Liu S, Cheng Y, Xu W, Bian Z. Protective effects of follicle-stimulating hormone inhibitor on alveolar bone loss resulting from experimental periapical lesions in ovariectomized rats. J Endod. 2010; 36:658–663.

26. Xiong J, Kang SS, Wang Z, et al. FSH blockade improves cognition in mice with alzheimer’s disease. Nature. 2022; 603:470–476.

27. Greendale GA, Huang MH, Wight RG, et al. Effects of the menopause transition and hormone use on cognitive performance in midlife women. Neurology. 2009; 72:1850–1857.

28. Greendale GA, Sowers M, Han W, et al. Bone mineral density loss in relation to the final menstrual period in a multiethnic cohort: Results from the Study of Women’s Health Across the Nation (SWAN). J Bone Miner Res. 2012; 27:111–118.

29. Greendale GA, Sternfeld B, Huang M, et al. Changes in body composition and weight during the menopause transition. JCI Insight. 2019; 4:

30. Cauley JA, Ruppert K, Lian Y, et al. Serum sex hormones and the risk of fracture across the menopausal transition: Study of Women’s Health Across the Nation. J Clin Endocrinol Metab. 2019; 104:2412–2418.

31. Kawai H, Furuhashi M, Suganuma N. Serum follicle-stimulating hormone level is a predictor of bone mineral density in patients with hormone replacement therapy. Arch Gynecol Obstet. 2004; 269:192–195.

32. Antonio L, Priskorn L, Olesen IA, Petersen JH, Vanderschueren D, Jorgensen N. High serum FSH is not a risk factor for low bone mineral density in infertile men. Bone. 2020; 136:115366.

33. Juel Mortensen L, Lorenzen M, Jorgensen N, et al. Possible link between FSH and RANKL release from adipocytes in men with impaired gonadal function including Klinefelter syndrome. Bone. 2019; 123:103–114.

34. Sowers M, Zheng H, Tomey K, et al. Changes in body composition in women over six years at midlife: Ovarian and chronological aging. J Clin Endocrinol Metab. 2007; 92:895–901.

35. Bowen JD, Malter AD, Sheppard L, et al. Predictors of mortality in patients diagnosed with probable Alzheimer’s disease. Neurology. 1996; 47:433–439.

36. Meethal SV, Smith MA, Bowen RL, Atwood CS. The gonadotropin connection in Alzheimer’s disease. Endocrine. 2005; 26:317–326.

37. Ohlsson C, Nilsson ME, Tivesten A, et al. Comparisons of immunoassay and mass spectrometry measurements of serum estradiol levels and their influence on clinical association studies in men. J Clin Endocrinol Metab. 2013; 98:E1097–1102.

